# SENSITIVITY ANALYSIS OF A TUBERCULOSIS (TB) MATHEMATICAL MODEL

**DOI:** 10.1101/2022.05.01.22274549

**Authors:** Vegha Vaachia, Mande Timothy Terna

## Abstract

In this work, we have constructed a Mathematical model for the transmission dynamics of tuberculosis TB. Feasibility and positivity of solutions of the model are determined and it is established that the model is well posed and that the solutions are all positive. The disease-free equilibrium (DFE) is also determined and the basic reproduction number *R*_0_ is computed. Sensitivity analysis of the basic reproduction number is conducted to find parameters of the model that are most sensitive and should be targeted by intervention strategies. It was therefore, observed through sensitivity analysis that TB induced death (d) has a high impact on *R*_0_ and varies inversely with *R*_0_. Graphical simulation of the model parameters was performed. It was observed that model parameters such as infection rate (*β*), recruitment rate (*π*) and rate of movement from latent to active TB (*α*) are directly proportional to the basic reproduction number*R*_0_. Finally, it is observed that the basic reproduction number *R*_0_ is a decreasing function of the recovery rate (*γ*) and natural death rate (*μ*).

## 1 Introduction

According to World Health Organization Tuberculosis (TB) is an infectious disease caused by mycobacterium tuberculosis [20]. Tuberculosis is spread through the air from one person to another. The bacteria get into the air when someone who has a tuberculosis lung infection coughs, sneezes, shouts, or spits. People who are nearby can then possibly breathe the bacteria into their lungs and become infected [20]. Even though the disease is airborne, it is believed that TB is not highly infectious and so, occasional contacts with infectious person rarely led to infection. TB cannot be spread through handshakes, sitting on toilet seats or sharing dishes and utensils with someone who has TB [3].

Most TB infections will result as a latent infection where the body is able to fight the bacteria and stopping them from growing. The bacteria thus will become dormant and remain in the body without causing symptoms. However, when the immune system of a patient with dormant TB is weakened, the TB can become active and cause infection in the lungs or other parts of the body. Only those with active TB can spread the disease [3]. Symptoms of TB disease depend on where in the body the TB bacteria grow. Active TB cases may be pulmonary where it affects the lungs. The early symptoms usually include fatigue or weakness, unexplained weight loss, fever, chills, loss of appetite and night sweats. Since the symptoms are very much similar to a common cold people tend to treat it as one. When the infection in the lung worsens, it may cause chest pain, bad cough that last through weeks or longer and coughing up of sputum and/or blood [3,7]. There are also cases where the infection spreads beyond the lungs to other parts of the body such as the bones and joints, the digestive system, the bladder and reproductive system and the nervous system. This is known as extra pulmonary TB and the Symptoms will depend upon the organs involved. It is more common in people with weaker immune systems, particularly those with HIV infection [20,17] lungs [17].

Tuberculosis is treated by killing the bacteria using antibiotics. The treatment usually last at least six months in duration and sometimes longer up to twenty-four months. It involves different antibiotics to increase the effectiveness while preventing the bacteria from becoming resistant to the medicines. The most common medicines used for active tuberculosis are Isoniazid (INH), Rifampin (RIF), Ethambunol and Pyrazinamide [17]. People with latent tuberculosis are usually treated using a single antibiotic to prevent them from progressing to active TB disease later in life. The only currently available vaccine is Bacillus Chalmette–Guerin (BCG) A number of new vaccines are currently in development [17].

TB is one of the top 10 causes of death worldwide. In 2015, 10.4 million people fell ill with TB, and 1.8 million died from the disease including 0.4 million among people with HIV [20]. Over 95% deaths occur in low and middle-income countries. Six countries account for 60% of the total, with India leading the count, followed by Indonesia, China, Nigeria, Pakistan and South Africa. In 2015, an estimated 1 million children became ill with TB and 170 000 died of TB excluding children with HIV.TB is also the leading killer of HIV positive people. In 2015, 35% of HIV deaths were due to TB. Globally in 2015, 480 000 people developed multi drug –resistant TB (MDR-TB) [20].

It is estimated that 2.7 million out of the 9.6 million infected people live in Africa [19]. Nigeria as a nation is ranked 10th among the 22 high-burden TB countries in the world. WHO estimates 210,000 new cases of all forms of TB occurred in the country in 2010, equivalent to 133/100,000 population. There were an estimated 320,000 prevalent cases of TB in 2010, equivalent to 199/100,000 cases [18].

## 2 Literature Review

In this section, we undertake a review of previous works carried out to study the transmission dynamics of tuberculosis which provide the perspective of our work. In 2013, [13] formulated an SEIR model with the assumption of permanent immunity and constant recruitment of infected individuals into the population. There was no disease-free equilibrium due to the constant recruitment of infected individuals. The model was used to analyze the local stability of the endemic equilibrium. A numerical solution was presented and this showed the existence of a globally asymptotically stable endemic equilibrium under certain parameter restrictions.

In 2011, [6] presented an SLIR model, the aim of the research was to determine the effect of stress on TB dynamics and treatment under the assumption that recovered individuals gets re-infected. It was concluded that as long as there are no interventions (treatment and health education campaigns) to control the spread of the disease, the disease will not be wiped out from the population as well as stress. It was further noted that there is a direct relationship between stress and the reproduction numbers, that is, as the rate of secondary infective increase also the rate of stress increases. Also, when treatment and Health education campaigns is given to infected individuals, stress can completely be wiped out of the population and thus reducing the rate of infection which increases the recovery rate in all classes.

In 2013, [5] examined in detail, a mathematical model for the transmission and prevalence of tuberculosis and the solution using differential equations. The following assumptions were made before the model was formulated. That, the population has a constant size of N, where birth and death occur at equal rates and that the newborns are susceptible (no inherited immunity). Also, there is no restriction of age, mobility or other social factors. It was also assumed that once infected with tuberculosis bacteria, you become exposed to the environment before becoming infectious. Based on the assumptions, an SEIR model was formulated. The model gave a basic reproductive number *R*_0_ = 1.09305 > 1. This means that the disease is endemic in the population and this is due to the high level of the transmission rate in the population.

In 2016, [2] in his paper titled “a mathematical analysis of HIV/TB co-infected” developed and analyzed a mathematical model of HIV/TB co infection. The model subdivides the human population into six compartments namely; Susceptible TB individuals, TB-infectious individuals, TB-recovered individuals, HIV-infected individuals, co-infected individuals and individuals with full blown AIDS. The stability analysis of the Disease-Free Equilibrium State (DFE) of the model shows that it will be stable if the population is sustainable. The analysis of the Endemic Equilibrium State (EES) shows that it is stable. This is in conformity with the real-life scenario. That is, whenever HIV is present, the patient may likely be infected with TB if proper and timely care is not given. Also, early detection of HIV and TB cases and provision of early treatment can help to control the disease.

In 2012, [14] developed an SEI model; the aim was to determine the existence of solution of a tuberculosis model. It was shown through the existence theorem that there exists a unique solution of the model.

This work therefore is an extension of the work of [14] by including the recovered compartment and taking into consideration the fact that individuals who recover from TB acquire temporary immunity and can become susceptible after recovery. It is therefore an SLIRS model.

## 3 Model Formulation

The model subdivides the total human population into Susceptible (S), Latent (L), Infectious (I) and Recovered individuals (R) so that that N= S+L+I+R as shown in table 1.

**Table 1:** Variables of the model.

| S/No. | Variable/Parameter | Description |
| --- | --- | --- |
| 1 | $S$ | Susceptible individuals |
| 2 | $L$ | Latent individuals |
| 3 | $I$ | Infectious individuals |
| 4 | $R$ | Recovered individuals |

The derivation of the model equations is represented diagrammatically in figure 1. The susceptible population consists of newly recruited individuals into the population by birth or immigration at a rate *π* and loss of immunity by recovered individuals at a rate *ω*. The population decreases as a result infection due to contact with infectious individuals at a rate *β* and natural death at a rate *μ*.

**Figure1:**
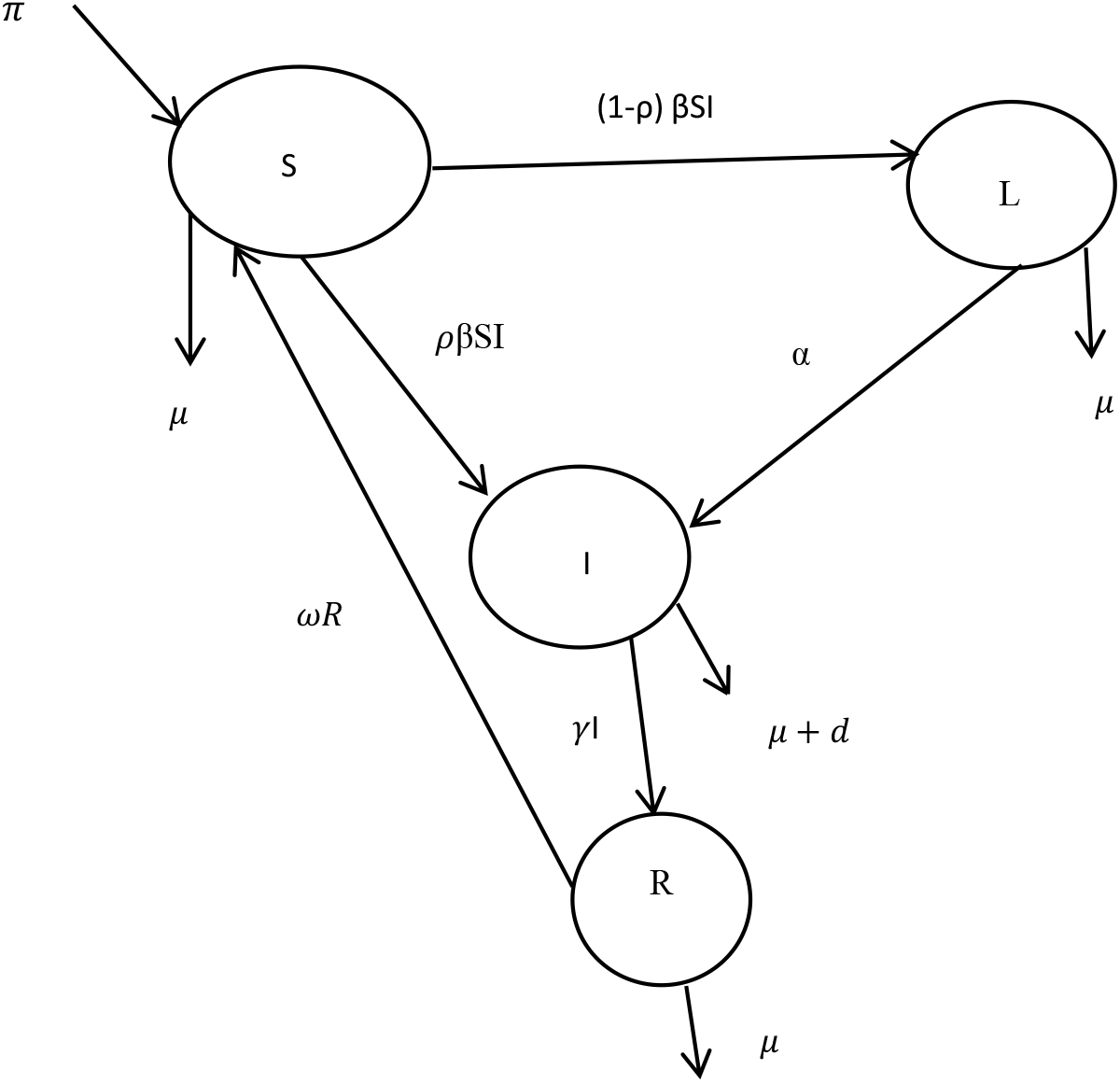
Schematic diagram for the model.

The population of latent individuals consists of a proportion of infected individuals who progresses slowly at a rate (1-*ρ*), the population declines as a result of natural death at a rate *μ*.

The population of infectious individuals consist of a proportion of newly infected individuals who progresses fast at rate *ρ* and individuals from the latent state at a rate *α*. The population reduces due to natural death at a rate *μ* and TB induced death at a rate d.

The Population of recovered individuals increases following recovery of infectious individuals at a rate *γ*. The population declines due to natural death at a rate *μ* and loss of immunity at a rate *ω*.

The model is governed by the following equations:

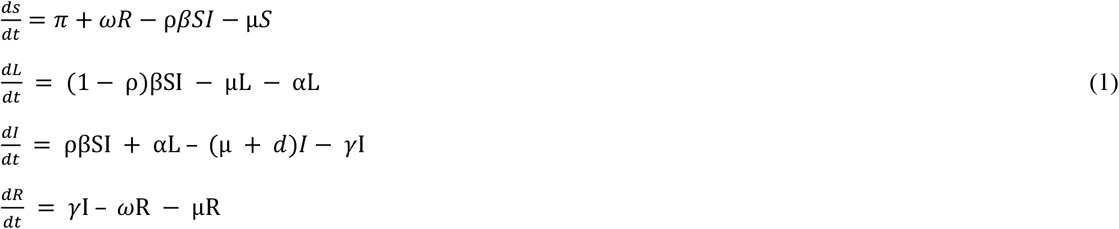

With the initial condition *S*(0) = *S*_0_, L(0) = *L*_0_, I(0) = *I*_0_, R(0) = *R*_0_ *and N*(0) = *N*_0_.

*N* = *S* + *L* + *I* + *R*, is the total population size changing at the rate

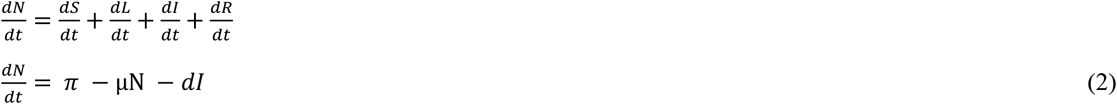

## 4 Basic Properties of the Model

### 4.1 Feasibility of the model

#### Lemma

The solutions of the system (1) are feasible for all *t* > 0 if they enter the invariant region Ω.

#### Proof

Let,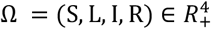 be any solution of the system (1) with non-negative initial conditions. From equation (2) in the absence of the disease (TB), *d* = 0 and (2) becomes:

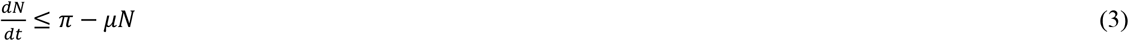

Integrating (2) and using the initial condition *N*(0) = *N*_0_, we obtain

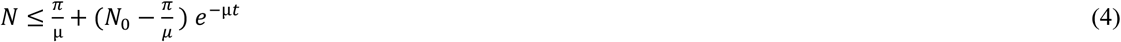

As 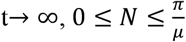

The total population approaches 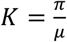 as *t* → ∞ which is commonly termed carrying capacity. Therefore, the feasible region for the model is 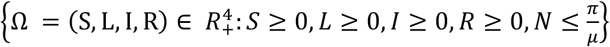

In this region, the model (1) is biologically feasible. Here whenever 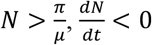, which means the population reduces asymptotically to the carrying capacity and whenever 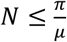, every solution with initial condition in Ω remains in that region for *t* > 0, so that the model is well posed in Ω. Therefore, the region Ω is positively invariant (that is the solution remains positive for all time t) and the model is well posed and biologically meaningful.

### 4.2 Positivity of Solutions

#### Lemma

Let the initial data be 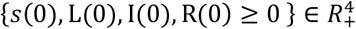. Then the solution set {*s*(t), L(t), I(t), R(t)} of the system (1) is positive for all t >0.

#### Proof

From equation (1), we have

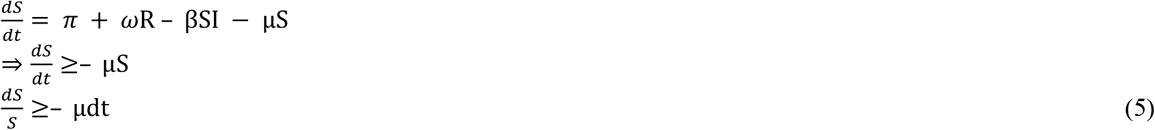

Integrating (5), we get S(t) ≥ S(0)*e*^*∫*–μdt^≥ 0, since μ ≥ 0

Similarly:

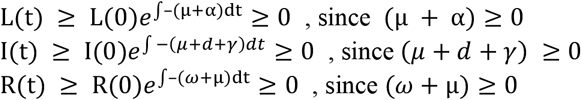

Thus, the solution set {*s*(t), L(t), I(t), R(t)} of the system (1) is positive for all t >0.

## 5 The Disease-Free Equilibrium (DFE)

The disease-free equilibrium (DFE) of a disease model is its steady state solutions in the absence of infection or disease. We denote the DFE by *E*_0_. At the disease-free equilibrium,

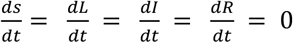

Let *E*_0_ = {*s*_0_, *L*_0_, *I*_0_, *R*_0_} be the equilibrium point of the model (1), in the absence of TB infection, *L*_0_ = 0, *I*_0_ = 0. Thus 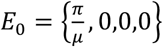 represents the state of the system (1) in which there is no infection.

## 6 The Basic Reproduction Number

The basic reproduction number denoted by R_0_ is the expected number of secondary TB infections produced in a completely susceptible population by a typical infective individual. It is one of the most useful threshold parameters in mathematical epidemiology [1].

If R_0_ < 1, this implies that on average an infected individual produces less than one new infection during the infectious period and the disease dies out. Conversely, if R_0_ > 1, then each infectious individual produces on average more than one new infection and disease spreads in the population [1].

For a single infected compartment, *R*_0_ is simply the product of the infection rate and the mean duration of the infection. But for complicated models, this simple definition of *R*_0_ is insufficient. We therefore compute the basic reproduction number *R*_0_ using the next generation method as follows [8].

The associated matrices *F*_*i*_ and *V*_*i*_ for new infection in the infected compartments and the remaining transfer terms are given respectively by

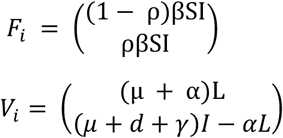

Next, we evaluate F and V which are the Jacobean of *F*_*i*_ and *V*_*i*_ respectively at *E*_0_ such that F is non-negative and V is a non-singular matrix. We denote the Jacobean of *F*_*i*_ and *V*_*i*_ by *J*(*F*_*i*_) and *J*(*V*_*i*_) respectively. Thus,

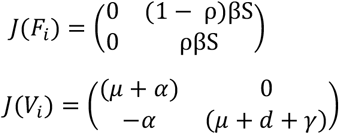

F = *J*(*F*_*i*_) at *E*_0_ and V = *J*(*V*_*i*_) at *E*_0_, Thus,

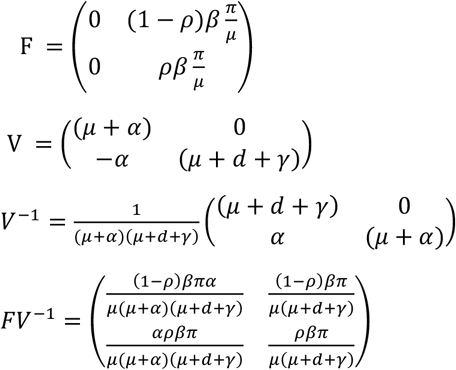

We next compute |*FV*^−1^− *λI*| = 0 as follows:

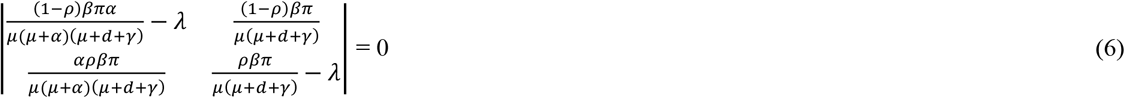

Let 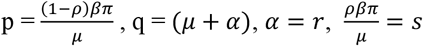 and (*μ* + *d* + *γ*) = *p*, then, equation can be written as:

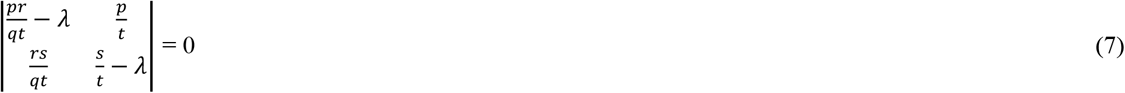

The eigen values of (7) are

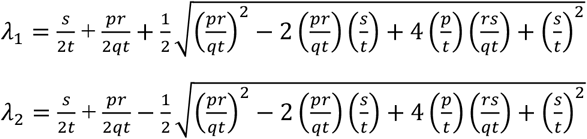

The basic reproduction number *R*_0_ is the spectra radius (dominant eigenvalue) of the matrix *FV*^−1^. Thus,

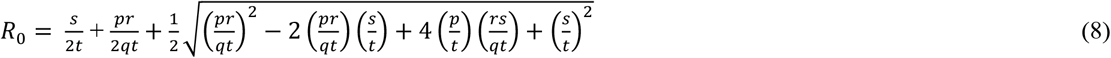

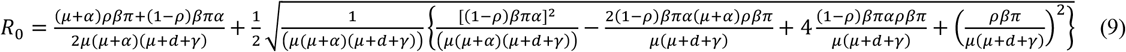

It can be shown that when *R*_0_ < 1, the disease-free equilibrium is locally asymptotically stable, otherwise it is unstable.

## 7 Sensitivity Analysis

Sensitivity analysis is used to determine how “sensitive” a model is to changes in the value of the parameters of the model and to changes in the structure of the model. Sensitivity analysis helps to build confidence in the model by studying the uncertainties that are often associated with Parameter in the model [15].

Sensitivity indices allow us to measure the relative change in a state variable when a parameter changes. Sensitivity analysis is commonly used to determine the robustness of model predictions to parameter values (since there are usually errors in data collection and presumed parameter values). Thus, we use it to discover parameters that have a high impact on *R*_0_ and should be targeted by intervention strategies [15].

If the result is negative, then the relationship between the parameters and *R*_0_ is inversely proportional. In this case, we will take the modulus of the sensitivity index so that we can deduce the size of the effect of changing that parameter. On the other hand, a positive sensitivity index means *R*_0_ varies directly with the parameter [15]. The explicit expression of *R*_0_ is given by the equation (9). Since *R*_0_ depends only on seven parameters *β,π, ρ, d, γ, α* and *μ*. we derive an analytical expression for its sensitivity to each parameter using the normalized forward sensitivity index as follows [8].

A small perturbation *δτ* of a parameter *τ* and the corresponding change in *R*_0_ as *δR*_0_ is given by

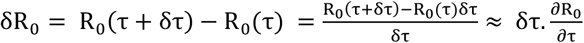

The normalized sensitivity index 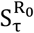 is d defined as 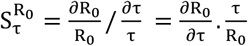

Thus, the normalized sensitivity indices for the seven parameters are obtained using the values in table 2. Since most of the parameter values were not readily available, we use data from literature and the missing data were estimated.

**Table2:** Parameter values, sources and sensitivity indices.

| S/NO | Parameter | Description | Value | Source | Sensitivity Index |
| --- | --- | --- | --- | --- | --- |
| 1 | $\beta$ | Infection rate | 3.0000 | CDC | +1 |
| 2 | $\pi$ | Recruitment rate | 0.0001 | Estimated | +1 |
| 3 | $\rho$ | Rate of fast progression | 0.8500 | Estimated | 0.0308 |
| 4 | $d$ | TB induced death rate | 0.4605 | American journal of epidemiology | -0.8145 |
| 5 | $\gamma$ | Recovery rate | 0.0862 | American journal of Epidemiology | -0.1525 |
| 6 | $\alpha$ | Latent to infectious progression rate | 0.5000 | Estimated | 0.0052 |
| 7 | $\mu$ | Natural death rate | 0.0187 | PLOS Journal | -1.0383 |

From table 2, we observe that the values of 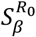 and 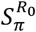 are exactly +1. This means that an increase in *β* and *π* will lead to an increase in *R*_0_ in the same proportion. Similarly, a decrease in *β* and *π* will cause *R*_0_ to decrease in the same proportion since they are directly proportional. We also observe that 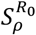 and 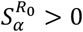 hence the parameters *ρ* and *α* are directly proportional to *R*_0_. Furthermore, 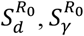 and 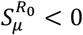, these means that the parameters *d, γ* and *μ* are inversely proportional to *R*_0_

## 8 Numerical Simulations

## 9 Discussion

In this section, we undertake a discussion of the results of figures under numerical simulations. All the figures were generated from equation (6) with the help of MATLAB 2016 using parameter values in table 2.

Figure2 shows a direct relationship between the basic reproduction number (*R*_0_) and the infection rate (*β*). The values of (*β*) were varied as shown on the figure. The result agrees with reality in the sense that as the rate of infection of TB increases, the number of individuals that will be infected in the population also increases there by increasing the basic reproduction number.

**Figure2:**
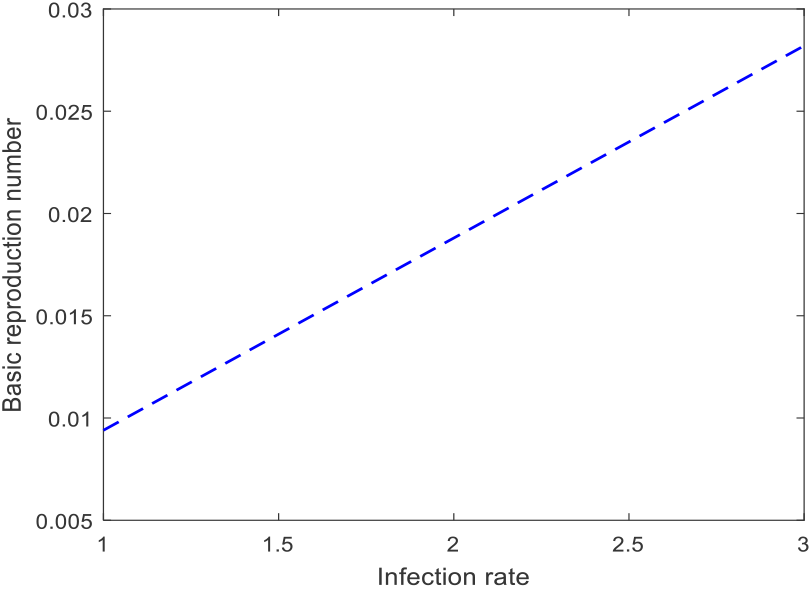
Effect of infection rate (*β*) on the basic basic reproduction number (*R*_0_)

Figure3 shows that the basic reproduction number (*R*_0_) is an increasing function of of the recruitment rate (*π*). Here the values of the recruitment rate (*π*) were varied as shown on the figure. This relationship is in line with real life scenario because if children are born without vaccination at birth, the more their chances of being infected with TB and thus infect others.

**Figure3:**
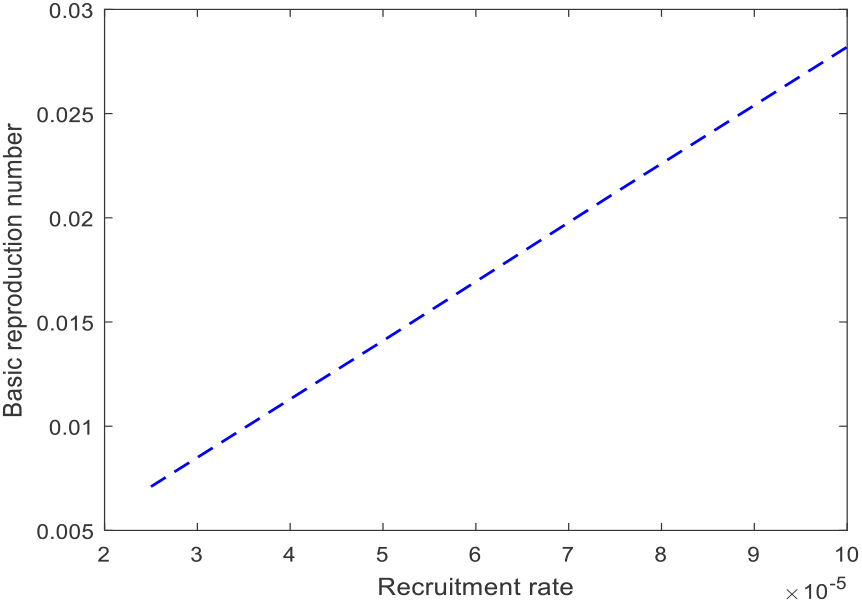
Effect of recruitment (*π*) on the reproduction number (*R*_0_)

In figure4, the basic reproduction number (*R*_0_) is a decreasing function of TB induced death d. Values of d were varied as shown on the figure. This also agrees with reality because as more infectious individuals die as a result of TB infection, the infection rate will also be reduced.

**Figure4:**
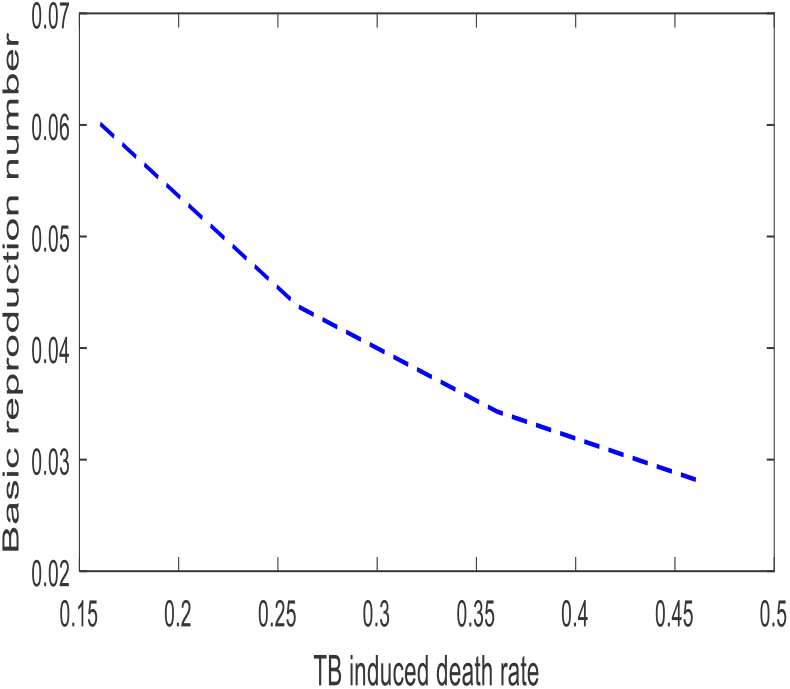
Effect of TB induced death Rate (d) on the basic reproduction number (*R*_0_)

Figure 5 shows an inverse relationship between the basic reproduction number (*R*_0_) and the recovery rate *γ*.This is true because if proper treatment is given to infective individuals, less people will be infected there by reducing the basic reproduction number.

**Figure5:**
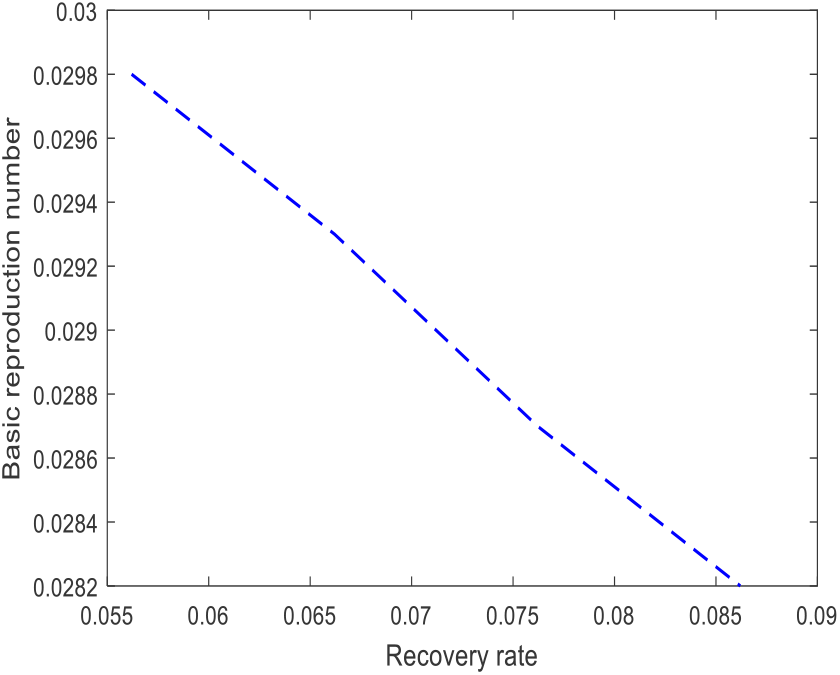
Effect of recovery Rate (*γ*) on the basic reproduction number (*R*_0_)

In figure 6, the basic reproduction number (*R*_0_) varies directly with rate of progression from latent to active TB (*α*). Values of (*α*) were varied as shown on the figure. The relationship also agrees with reality in that if latently infected individuals are not subjected to early treatment, they will become infectious and hence cause more infections.

**Figure 6:**
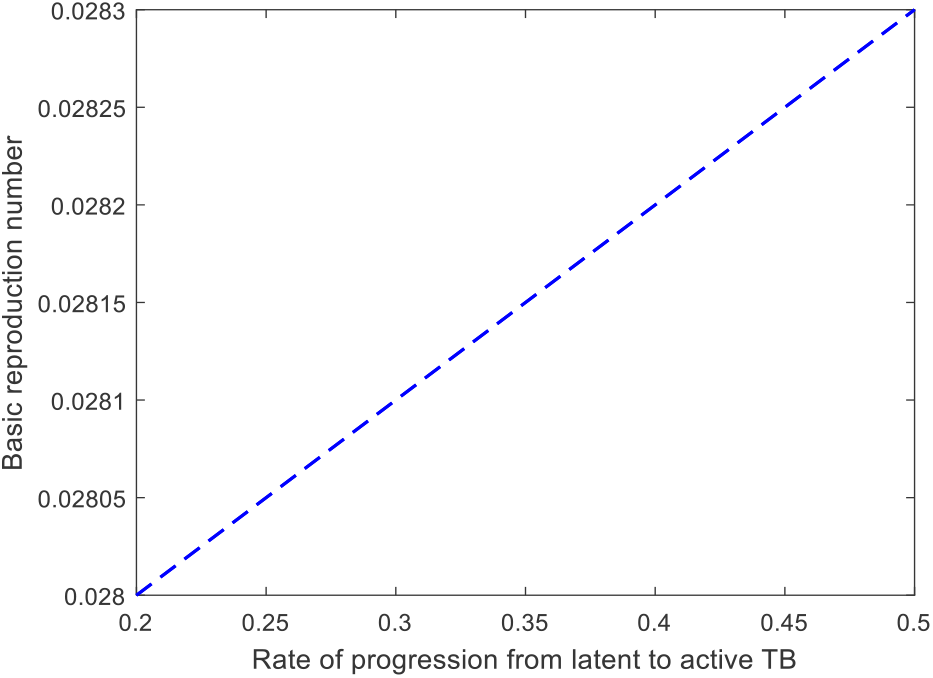
Effect of Rate of progression from latent to active TB (*α*) on the basic reproduction number (*R*_0_)

Finally, in figure7, we have seen that by increasing the values of the population natural death rate *μ*, as shown on figure 6, the basic reproduction number *R*_0_ is a decrease. This is also in line with reality because as more people die naturally, there will be fewer people to be infected with TB.

**Figure7:**
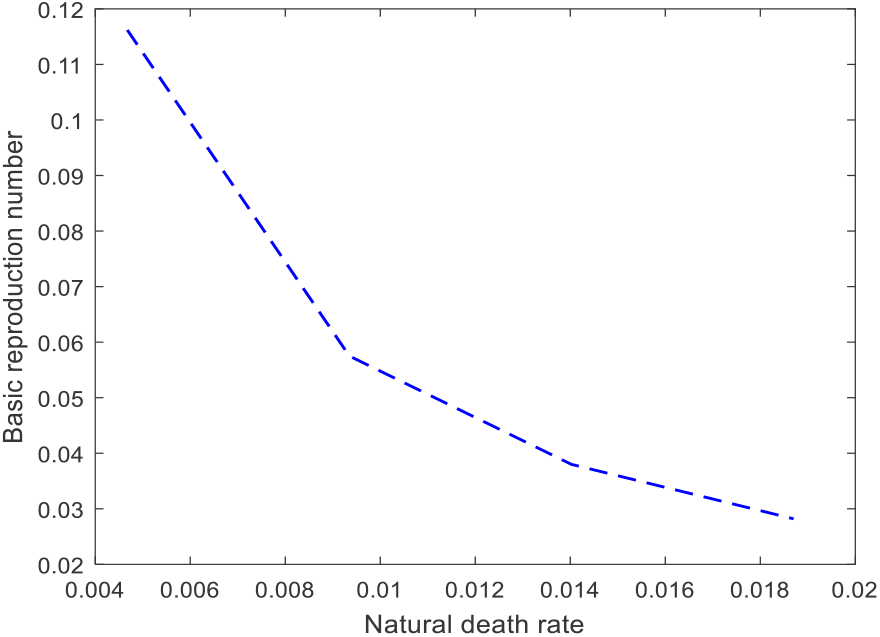
Effect of natural death rate (*μ*) on the basic reproduction number (*R*_0_)

## 10 Conclusion

In this work, we have constructed a Mathematical model for the transmission dynamics of tuberculosis TB. Feasibility and positivity of solutions of the model were determined and it was established that the model is well posed and that the solutions were all positive. The disease-free equilibrium state of the model was also determined as well as the basic reproduction number *R*_0_.

Sensitivity analysis of the basic reproduction number and graphical simulation of the model parameters were performed. It was observed that model parameters such as infection rate (*β*), recruitment rate (*π*) and rate of movement from latent to active TB (*α*) are directly proportional to the basic reproduction number *R*_0_. Thus to reduce the rate of spread of TB in the population, the study therefore recommends that these parameters be targeted by way of intervention strategies. For example, vaccination of children at birth, education campaign and early treatment of infected individuals can help reduce the spread of the disease. It was observed through sensitivity analysis that TB induced death (d) has a high impact on *R*_0_ and varies inversely with *R*_0_. The study recommends that infectious individuals who are capable of spreading the disease and are at the risk of dying due to infection should also be given proper treatment.

Finally, it is observed that the basic reproduction number *R*_0_ is a decreasing function of the recovery rate (*γ*) and natural death rate (*μ*). This means if proper treatment is given to infectious individuals, there will be an increased recovery rate and decreased TB infection and thus most of the deaths in the population will be natural.

## Data Availability

All data is contained in the manuscript and supporting files

https:www

